# *ABCA4-*Related Retinopathies in Lebanon: a novel mutation and significant heterogeneity

**DOI:** 10.1101/2023.11.09.23298241

**Authors:** Mariam Ibrahim, Lama Jaffal, Alexandre Assi, Charles Helou, Said El Shamieh

## Abstract

Mutations in *ATP-binding cassette transporter type A4* (*ABCA4*) have been linked to several forms of inherited retinal diseases (IRDs) besides the classically defined Stargardt disease (STGD), known as *ABCA4* retinopathies. *ABCA4* is a sizeable locus harboring 50 exons; thus, its analysis has revealed a rich area of genetic information comprising at least 1,200 disease-causing mutations of varied severity and types. Due to the clinical and genetic heterogeneity, diagnosing *ABCA4* retinopathies is challenging. To date, no *ABCA4*-retinopathy has been detected in Lebanon. Using next-generation sequencing, we sought to pinpoint the mutation spectrum in seven families with different forms of IRDs: STGD, rod-cone and cone-rod dystrophies (RCD and CRD, respectively). Eight *ABCA4* mutations were found, including one novel; c.4330G>C; p.(Trp1408Cys). Three families were diagnosed with CRD, two with STGD, and two others with RCD. In conclusion, our study revealed a novel ABCA4 mutation and showed significant genotypic and phenotypic heterogeneity in Lebanon.

## Introduction

Inherited retinal dystrophies (IRDs) are a set of monogenic disorders marked by photoreceptor degeneration or impairment [1, 2]. These diseases are well defined by a high degree of clinical and genetic variation, with over 270 genes implicated [1]. Interestingly, the age of onset, the progression rate, manifestation with extra-ocular symptoms, and the etiological gene may assist in classifying IRDs into more than 20 distinct phenotypes [3, 4]. The most prevalent form of IRDs is rod-cone dystrophy (RCD; MIM 613862), which affects over one million individuals worldwide and is defined by the primary death of rods subsequently accompanied by secondary deterioration of cone photoreceptors [3, 5]. When cone photoreceptor degeneration occurs initially, followed by rod dysfunction in later stages, this is called cone-rod dystrophy (CRD; MIM 601777), represented by progressive degeneration and loss of the central retina [6, 7]. Other aspects of IRDs that appear with central vision loss include macular dystrophies (MD) that affect mainly the macula [3, 7]. With an incidence rate of 1 in 8,000–10,000 individuals, Stargardt disease (STGD; MIM 248200) emerges as the prevailing aspect of MD with an autosomal recessive mode of inheritance affiliated with etiological mutations in the ATP-binding cassette transporter type A4 (*ABCA4*) [8].

Since the 1997 identification of the *ABCA4* gene by Allikmets and colleagues, more than 1,200 distinct disease-causing mutations of varying severity have been reported [9–11]. *ABCA4* variations have been associated with phenotypes other than the commonly recognized STGD, including fundus flavimaculatus, age-related MD, and some forms of CRD and RCD [9, 10]. The *ABCA4* locus shows significant disparities in disease-causing alleles across racial and ethnic groups and exhibits founder mutations in different populations [11]. Considering the massive clinical and genetic heterogeneity, an accurate and thorough molecular diagnosis of *ABCA4*-associated retinopathies is critical [12]. However, the allelic heterogeneity of *ABCA4* has made genetic analyses of these gene-associated IRDs very challenging [13]. Direct Sanger sequencing of all *ABCA4* exons (50 exons) has uncovered between 60%-80% of the pathogenic alleles [12]. Notably, next-generation sequencing (NGS) platforms have proven to find novel *ABCA4* mutations, demonstrating their ability as more comprehensive approaches for systematic genetic screening of large cohorts [14, 15]. Presently, NGS is critical for obtaining a prompt and precise genetic diagnosis, which is required to provide patients and their families with the appropriate genetic counseling [1, 16]. The relevance of genetic diagnosis through the implementation of comprehensive and affordable sequencing technologies lies in identifying the disease-causing mutations that may result in finding the phenotype-genotype correlations, establishing a clear interpretation of the pathophysiological mechanisms, and tailoring the approach of personalized therapy [5]. Many *ABCA4* mutations associated with different forms of IRDs have been reported worldwide, but none in Lebanon. Herein, the aim was to detect causative *ABCA4* mutations in Lebanese patients from seven families diagnosed with varying forms of IRDs.

## Materials and Methods

### Ethical considerations and Clinical Examinations

All our procedures adhered to the tenets of the Declaration of Helsinki. Beirut Arab University’s institutional review board under the IRB number 2017 H-0030-HS-R-0208 granted ethical approval. All our participants were recruited at Beirut Eye and ENT Specialist Hospital (Lebanon) where they received carried out a clinical ophthalmologic assessment and provided informed written consent before participation, as previously described [17].

### Mutational Screening

All our participants gave whole blood samples. Genomic DNA extraction was done by Qiagen’s QIAamp DNA Mini Kit (Hilden, Germany). Whole exome sequencing (WES) was executed to inspect the DNA samples of all the indexes except F3:I3 and F9:I9.1, sequenced through targeted NGS as described previously [15]. Common polymorphisms that had a minor allele frequency (MAF) greater than 0.01 were all omitted using various public databases, including GnomAD (https://gnomad.broadinstitute.org/) [18]. This step was followed by annotation type-based filtration, where we removed in-frame Indels, intronic mutations, synonymous mutations, and variations in untranslated regions. Contrariwise, priority was given to mutations in exons or splice sites that resulted in nonsense, missense variations, or frameshift Indels. Next, we checked whether the candidate mutations were reported as homozygous in GnomAD.

### Pathogenicity assessment of the Candidate Mutations

The conservation of substituted amino acids in various species, such as primates and main placental mammals, was checked using the University of California at Santa Cruz (UCSC) genome browser [19]. Information regarding the details was previously described [20]. The possible effect of the amino acid substitution was assessed by scale-invariant feature transform (SIFT) [21], Polymorphism Phenotyping v2 (PolyPhen-2) [22], and MutationTaster2 [23]. Several public databases were utilized to determine if the candidate mutation causing IRD was previously known [24], [25].

### Segregation Analysis

The candidate mutations detected by NGS were amplified by conventional polymerase chain reaction (T100, Biorad, Kaki Bukit, Singapore) and then validated (Applied Biosystems 3730xl DNA Sequencer, Courtaboeuf, Les Ulis, France) to exclude the possibility of false positive results. For segregation purpose, unidirectional Sanger sequencing was applied to all available family members’ DNA.

## Results

### Ophthalmic data

The current study involves seven Lebanese families, each having at least one affected member. In family 3, index F3:I3 diagnosed with STGD in their 20s. Parents of F3:I3 are somehow relatives but not first cousins (Figure 1). Color fundus photographs of F3:I3 revealed mild pigmentary changes in the posterior pole and outside the vascular arcades (Figure 2a). Fluorescein angiography showed granular hyperfluorescence in the posterior pole with focal hyperfluorescence at the macula (Figure 2b). Additionally, optical coherence tomography (OCT) exhibited diffuse thinning of the retinal layers (Figure 2c). Electrooculogram (EOG) demonstrated no light rise and a subnormal Arden ratio of 1.68 on the right eye and a reduced Arden ratio of 1.51 on the left eye, which are below the normal value (>1.8); electroretinogram (ERG) revealed reduced scotopic and photopic responses (data not shown).

**Figure 1.**
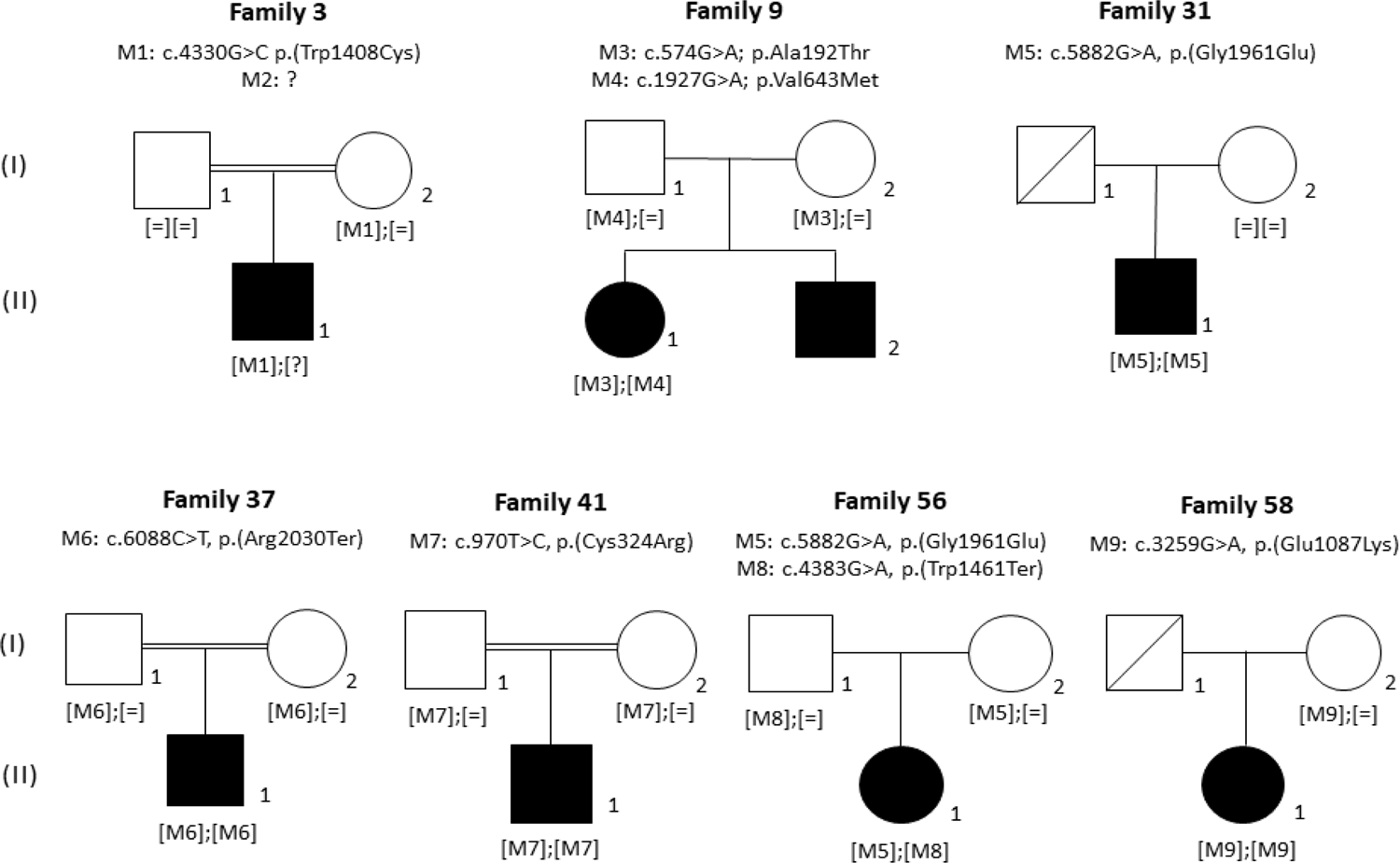
Pedigrees of seven families with mutations in *ABCA4* gene. White symbols represent members who are unaffected. Symbols in black denote affected individuals. Males and females are represented by square and round symbols, respectively. Individuals who have died are denoted by a slash. Double horizontal lines represent consanguineous marriages. M: mutation.

**Figure 2:**
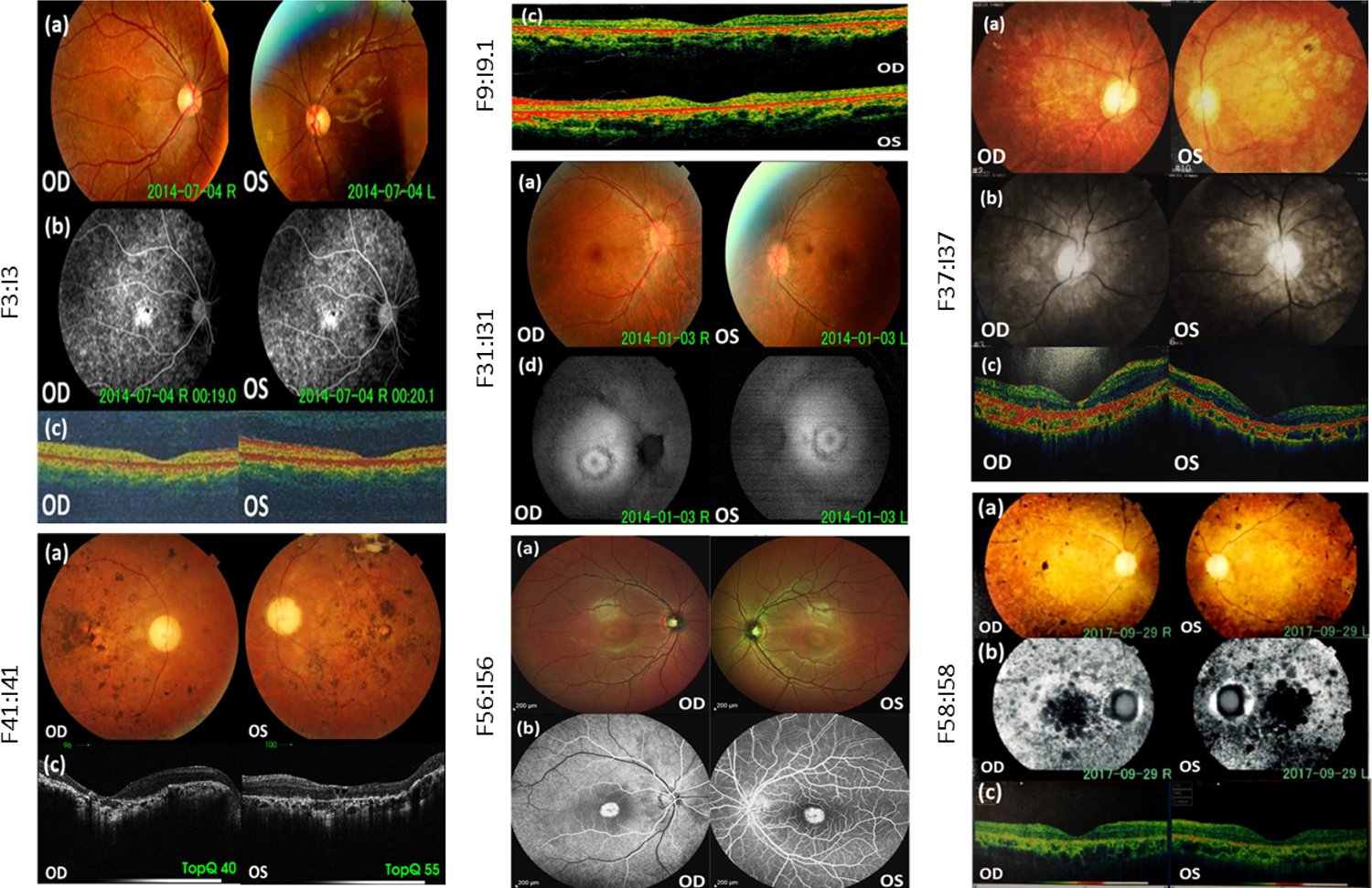
Color fundus photographs (a), fluorescein angiography (b), optical coherence tomography (OCT) scans (c), and autofluorescence pictures (d) of patients F3:I3, F9:I9.1, F31:I31, F37:I37,F41:I41, F56:I56 and F58:I58. OD = oculus Dexter; OS: ocular sinister.

The index of family 9 does not have any previous cases in her family. In her 20s, she received a diagnosis of CRD, and her OCT revealed thinning of the retinal layers at the macula in both eyes (Figure 2c). ERG demonstrated normal photopic and very reduced scotopic responses (data not shown). Moreover, EOG displayed a normal Arden ratio of 2 on the right eye and a significantly subnormal Arden ratio of 1.6 on the left eye.

F31:I31, diagnosed with RCD in their 20s with no known family history. Color fundus photography of this patient showed mild pigmentary changes in the posterior pole and outside the vascular arcades (Figure 2a). His autofluorescence examination showed increased hyperfluorescence at the macula (Figure 2d). Besides, the ERG of this individual demonstrated reduced photopic and significantly reduced scotopic responses. Additionally, F31:I31 exhibited reduced EOG with an Arden ratio of 1.29 on the right eye and 1.25 on the left eye (data not shown).

Index F37:I37, issued from a consanguineous marriage, and was diagnosed with CRD in their 30s. Clinical findings of this patient indicated reduced photopic and scotopic ERG responses. Moreover, the EOG of this patient exhibited a reduced Arden ratio of 1.25 and 1.24 on the right and left eyes, respectively (data not shown). Color photographs revealed optic disc parlor and atrophy in the posterior pole (Figure 2a). OCT scans showed thinning of the retinal layers and hyper-reflectivity at the level of the choroid (Figure 2c).

Family 41 exhibited consanguinity among parents, in which the father is affected (Figure 1). This family has an affected descendant, F41:I41. Color fundus examination showed marked pigmentary changes in the posterior pole with marked vascular attenuation and optic disc pallor (Figure 2a). Additionally, OCT demonstrated diffuse thinning of the retinal layers with cystic changes and focal scarring (Figure 2c). The ERG displayed a diminished photopic and scotopic responses. The clinical findings indicated a CRD.

Parents in family 56 are phenotypically normal but have an affected descendant, F56:I56, diagnosed with STGD in their 20s. Her color photograph showed an abnormal reflex at the macula (Figure 2a). Besides, fluorescein angiography revealed increased hyperfluorescence at the macula (Figure 2b). Clinical diagnosis of F56:I56 revealed macular dystrophy with relative preservation of macular function. ERG of this patient demonstrated slightly reduced photopic and scotopic responses (data not shown). Additionally, visual evoked potentials (VEP) was reduced for a small pattern.

Index F58:I58 has phenotypically non-affected parents, but she was diagnosed clinically with RCD. ERG revealed reduced photopic and very reduced scotopic (data not shown). Color photograph showed marked pigmentary changes in the posterior pole and outside the vascular arcades with marked vascular attenuation and optic disc pallor (Figure 2a). Fluorescein angiography demonstrated diffuse granular hyperfluorescence in the posterior pole with decreased fluorescence at the macula (Figure 2b). Additionally, OCT scan of the patient exhibited diffuse thinning of the retinal layers (Figure 2c).

### Genetic findings

We detected eight mutations in the *ABCA4* gene in seven Lebanese families (Table 2). For index F3:I3 of family 3, we found a mutation in the ABCA4 gene in heterozygosity state. However, the second mutated allele remains missing. The detected mono-allelic mutation is a missense mutation in exon 28 of the *ABCA4* gene: [M1]: c.4330G>C, p.(Trp1408Cys). M1 was not found in ExAC, gnomAD, or TopMed populations; at the protein level, it affects a conserved amino acid, Trp1408, with two exceptions. The substitution was also predicted to be damaging. Sanger sequencing validated the occurrence of M1 in a heterozygous state in the patient F3:I3 of this family (Figure S1 a). The mother was a heterozygous carrier of M1, while the father carried the wild-type allele. M1 is novel and has not been reported before in literature databases.

**Table 1.**
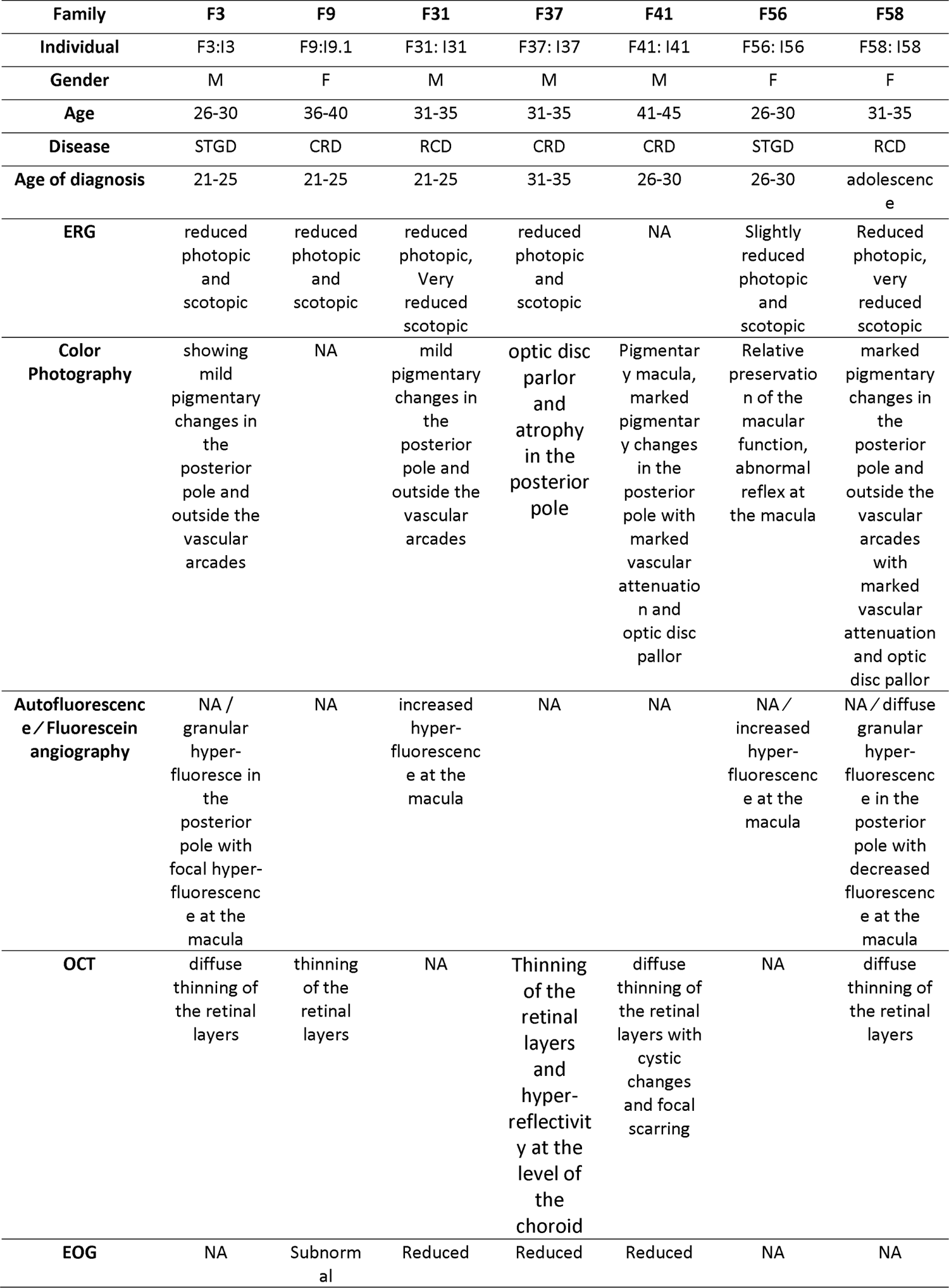
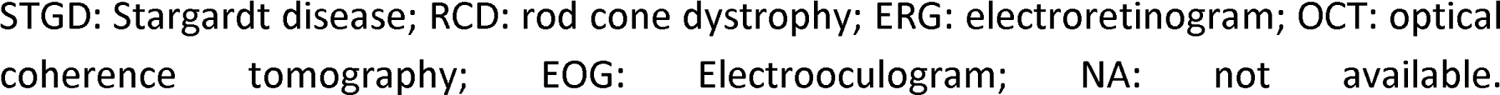
Clinical results identified in eight Lebanese patients with inherited retinal disorders.

| Family | F3 | F9 | F31 | F37 | F41 | F56 | F58 |
| --- | --- | --- | --- | --- | --- | --- | --- |
| Individual | F3: I3 | F9: I9.1 | F31: I31 | F37: I37 | F41: I41 | F56: I56 | F58: I58 |
| Gender | M | F | M | M | M | F | F |
| Age | 26-30 | 36-40 | 31-35 | 31-35 | 41-45 | 26-30 | 31-35 |
| Disease | STGD | CRD | RCD | CRD | CRD | STGD | RCD |
| Age of diagnosis | 21-25 | 21-25 | 21-25 | 31-35 | 26-30 | 26-30 | adolescence |
| ERG | reduced photopic and scotopic | reduced photopic and scotopic | reduced photopic, Very reduced scotopic | reduced photopic and scotopic | NA | Slightly reduced photopic and scotopic | Reduced photopic, very reduced scotopic |
| Color Photography | showing mild pigmentary changes in the posterior pole and outside the vascular arcades | NA | mild pigmentary changes in the posterior pole and outside the vascular arcades | optic disc pallor and atrophy in the posterior pole | Pigmentary macula, marked pigmentary changes in the posterior pole with marked vascular attenuation and optic disc pallor | Relative preservation of the macular function, abnormal reflex at the macula | marked pigmentary changes in the posterior pole and outside the vascular arcades with marked vascular attenuation and optic disc pallor |
| Autofluorescence/Fluorescein angiography | NA / granular hyper-fluoresce in the posterior pole with focal hyper-fluorescence at the macula | NA | increased hyper-fluorescence at the macula | NA | NA | NA / increased hyper-fluorescence at the macula | NA / diffuse granular hyper-fluorescence in the posterior pole with decreased fluorescence at the macula |
| OCT | diffuse thinning of the retinal layers | thinning of the retinal layers | NA | Thinning of the retinal layers and hyper-reflectivity at the level of the choroid | diffuse thinning of the retinal layers with cystic changes and focal scarring | NA | diffuse thinning of the retinal layers |
| EOG | NA | Subnormal | Reduced | Reduced | Reduced | NA | NA |

STGD: Stargardt disease; RCD: rod cone dystrophy; ERG: electroretinogram; OCT: optical coherence tomography; EOG: Electrooculogram; NA: not available.

**Table 2.**
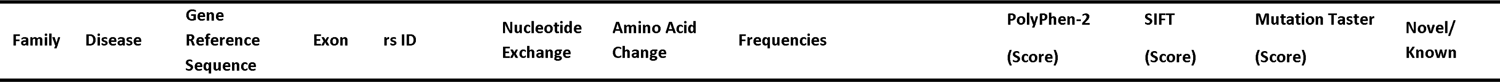
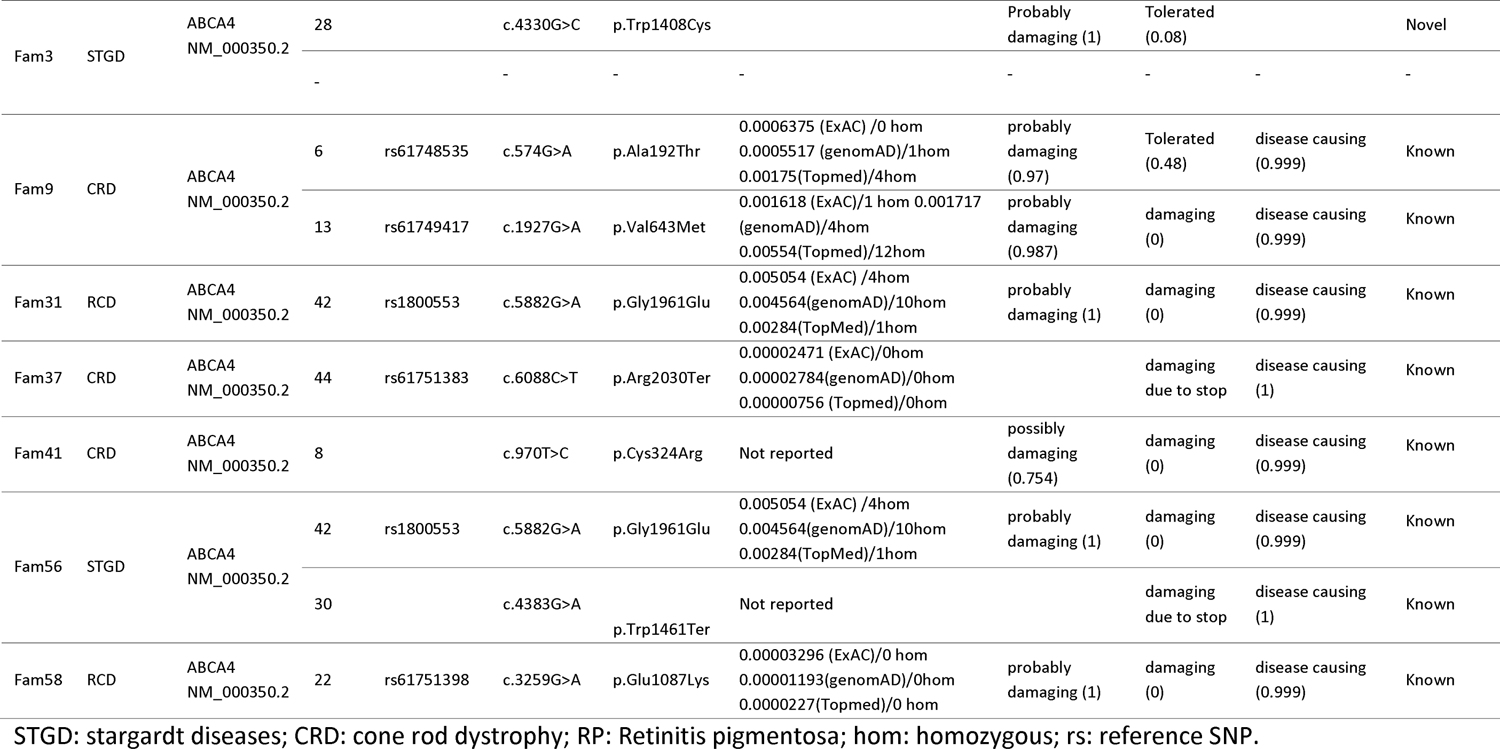
ABCA4 mutations in seven Lebanese families with different forms of inherited retinal diseases.

| Family | Disease | Gene<br>Reference<br>Sequence | Exon | rsID | Nucleotide<br>Exchange | Amino Acid<br>Change | Frequencies | PolyPhen-2<br>(Score) | SIFT<br>(Score) | Mutation Taster<br>(Score) | Novel/<br>Known |
| --- | --- | --- | --- | --- | --- | --- | --- | --- | --- | --- | --- |

|  |  |  |  |  |  |  |  |  |  |  |  |
| --- | --- | --- | --- | --- | --- | --- | --- | --- | --- | --- | --- |
| Fam3 | STGD | ABCA4<br>NM_000350.2 | 28 |  | c.4330G>C | p.Trp1408Cys |  | Probably<br>damaging (1) | Tolerated<br>(0.08) |  | Novel |
|  |  |  | - |  | - | - | - | - | - | - | - |
| Fam9 | CRD | ABCA4<br>NM_000350.2 | 6 | rs61748535 | c.574G>A | p.Ala192Thr | 0.0006375 (ExAC) /0 hom<br>0.0005517 (genomAD)/1hom<br>0.00175(Topmed)/4hom | probably<br>damaging<br>(0.97) | Tolerated<br>(0.48) | disease causing<br>(0.999) | Known |
|  |  |  | 13 | rs61749417 | c.1927G>A | p.Val643Met | 0.001618 (ExAC)/1 hom 0.001717<br>(genomAD)/4hom<br>0.00554(Topmed)/12hom | probably<br>damaging<br>(0.987) | damaging<br>(0) | disease causing<br>(0.999) | Known |
| Fam31 | RCD | ABCA4<br>NM_000350.2 | 42 | rs1800553 | c.5882G>A | p.Gly1961Glu | 0.005054 (ExAC) /4hom<br>0.004564(genomAD)/10hom<br>0.00284(TopMed)/1hom | probably<br>damaging (1) | damaging<br>(0) | disease causing<br>(0.999) | Known |
| Fam37 | CRD | ABCA4<br>NM_000350.2 | 44 | rs61751383 | c.6088C>T | p.Arg2030Ter | 0.00002471 (ExAC)/0hom<br>0.00002784(genomAD)/0hom<br>0.00000756 (Topmed)/0hom |  | damaging<br>due to stop | disease causing<br>(1) | Known |
| Fam41 | CRD | ABCA4<br>NM_000350.2 | 8 |  | c.970T>C | p.Cys324Arg | Not reported | possibly<br>damaging<br>(0.754) | damaging<br>(0) | disease causing<br>(0.999) | Known |
| Fam56 | STGD | ABCA4<br>NM_000350.2 | 42 | rs1800553 | c.5882G>A | p.Gly1961Glu | 0.005054 (ExAC) /4hom<br>0.004564(genomAD)/10hom<br>0.00284(TopMed)/1hom | probably<br>damaging (1) | damaging<br>(0) | disease causing<br>(0.999) | Known |
|  |  |  | 30 |  | c.4383G>A | p.Trp1461Ter | Not reported |  | damaging<br>due to stop | disease causing<br>(1) | Known |
| Fam58 | RCD | ABCA4<br>NM_000350.2 | 22 | rs61751398 | c.3259G>A | p.Glu1087Lys | 0.00003296 (ExAC)/0 hom<br>0.00001193(genomAD)/0hom<br>0.0000227(Topmed)/0 hom | probably<br>damaging (1) | damaging<br>(0) | disease causing<br>(0.999) | Known |

STGD: stargardt diseases; CRD: cone rod dystrophy; RP: Retinitis pigmentosa; hom: homozygous; rs: reference SNP.

For Index F9:I9.1 of family 9, NGS revealed a compound heterozygous mutation in *ABCA4;* two missense mutations [M3] [17]: c.574G>A; p.(Ala192Thr), rs61748535 in exon 6 and [M4]: c.1927G>A; p.(Val643Met), rs61749417 in exon 13. According to population databases, M3 is a rare mutation showing frequencies equal to 0.0006375, 0.0005517, and 0.00175 in ExAC, genomAD, and TOPMed databases, respectively, and found homozygous in several individuals in genomAD and TOPMed. Referring to the UCSC genome browser, it has been observed that M3 mutation impacts an amino acid residue (Ala192) that is conserved among multiple species, with the exception of four specific cases. Furthermore, based on PolyPhen-2 and MutationTaster algorithms, M3 is probably damaging and disease-causing. In contrast, SIFT reported it as tolerated. On the other hand, the M4 mutation appeared to be rare and homozygous in several individuals, as outlined in ExAC, GnomAD, and TOPMed population databases with respective frequencies of 0.001618, 0.001717, and 0.00554. M4 affects a conserved amino acid (Val643). Besides, based on the assessments of PolyPhen-2, SIFT, and MutationTaster, it was predicted that the M4 mutation is probably damaging, damaging and disease-causing, respectively. M3 [27–29] and M4 were known mutations [30, 31]. The existence of both mutations was verified by Sanger sequencing in F9:I9.1. The father and the mother of family 9 were heterozygous carriers of M4 and M3, respectively, confirming an adequate co-segregation (Figure S1 b).

The index of family 31, F31:I31, carries a homozygous missense mutation within exon 42 of *the ABCA4* gene. This mutation is well-known in exon 42, [M5]: c.5882G>A; p.(Gly1961Glu), rs1800553. M5 exhibited a rare occurrence and was observed to be homozygous in some individual from ExAC, gnomAD, and TOPmed population databases (respective frequencies= 0.005054, 0.004564, and 0.00284). This mutation affects a conserved residue (Gly1961) in different species, except for one. Furthermore, according to the predictions made by PolyPhen-2, SIFT, and MutationTaster, the M5 mutation was determined to be probably damaging, damaging, and disease-causing respectively. Sanger sequencing analysis validated the presence of this homozygous mutation in F31:I31 (Figure S1 c). The father of the affected patient was deceased; however, the mother was found homozygous WT.

The index F37:I37 of family 37 had a homozygous nonsense mutation in the exon 44 of *ABCA4*. This substitution mutation that resulted in the appearance of a stop codon is [M6]: c.6088C>T; p.(Arg2030Ter), rs61751383. According to population databases, M6 is a rare and heterozygous mutation with frequencies equal to 0.00002471, 0.00002784, and 0.00000756 in ExAC, genomAD and TOPMed, respectively, affecting a conserved amino acid with one exception based on UCSC genome browser. Furthermore, M6 is predicted to be disease-causing on MutationTaster and damaging on SIFT. This mutation was validated by Sanger sequencing in F37:I37. Moreover, parents were found to be heterozygous carriers of M6 (Figure S1 d). Referring to literature databases, M6 is not a novel mutation [32].

Index F41:I41 presented a homozygous mutation [M7]: c.970T>C; p.(Cys324Arg) in exon 8. The parents were heterozygous for M7 (Figure S1 e), which was extremely rare and absent in online databases. Prediction tools revealed it as disease-causing. Based on the UCSC genome browser, the impacted amino acid (Cys324) is conserved with two variations in two species. This mutation was previously reported in a Chinese population [33].

In family 56, F56:I56 harbors a compound heterozygous mutation. The first is the missense substitution in exon 42 [M5], while the second is a nonsense mutation in exon 30 [M8]: c.4383G>A; p.(Trp1461Ter) affecting a highly conserved amino acid. Notably, M8 was reported in the literature [34]. M8 was not shown in any population dataset, indicating that it is extremely rare. Moreover, M8 was shown to be disease-causing on MutationTaster and damaging on SIFT. Sanger sequencing validated the presence of M5 and M8 in F56:I56. Besides, it revealed that the disease co-segregated within the family, where the father was found heterozygous for M8 and the mother was heterozygous for M5 (Figure S1 f).

Mutational analysis in index F58:I58 revealed a homozygous missense mutation [M9] in exon 22. M9: c.3259G>A; p.(Glu1087Lys), rs61751398 is rare and never homozygous with an allele frequency of 0.00003296 in ExAC, 0.00001193 in genomAD and 0.0000227 in TOPMed. M9 is likely damaging on PolyPhen-2, damaging on SIFT, and disease-causing on MutationTaster. The affected amino acid (Gly1087) is also highly conserved, as found in the UCSC genome browser. The zygosity of the M9 mutation was verified through Sanger sequencing in the index patient. In contrast, M9 was heterozygous in the mother (Figure S1 g). This finding confirms the co-segregation of the mutation with the disease. The literature search showed M9 as a known variation [32].

## Discussion

To date, the analysis of the *ABCA4* gene has disclosed a bulk of genetic data with more than 1,200 mutations underlying IRDs of different severity and manifestations [11]. We found eight mutations in the *ABCA4* gene in a small Lebanese group, seven of which had been identified before in different populations, and one is novel, [M1]; c.4330G>C; p.(Trp1408Cys). The eight identified mutations were associated with varying forms of IRDs. Sanger sequencing verified all putatively pathogenic mutations, revealing their co-segregation with the associated phenotypes.

*ABCA4* is localized in 1p21–p22.1 on chromosome 1, consisting of 50 exons that encode a single-chain ATP-binding cassette transporter protein of 2,273 amino acids with a molecular weight of ∼250 kDa situated at the outer segments of rod and cone photoreceptors [9, 35]. ABCA4 protein moves all trans-retinal and toxic substances from the disc lumen to the photoreceptors’ cytoplasm. These by-products are transported to the retinal pigment epithelium (RPE) [36]. The association of bi-allelic mutations in *ABCA4* with different forms of IRDS comprises a loss-of-function component and photoreceptor stress due to faulty localization and folding of protein [35].

Determining the biallelic mutations may be challenging due to the *ABCA4*’s large size, the wide range of pathogenic variations it exhibits such as hypomorphic mutations, non-canonical splice site mutations, and lately, deep-intronic mutations [37]. In line with previous literature [38], targeted sequencing of the patient in family 3 revealed only one mutant allele, while the second mutation is still missing, thus rendering the case of family 3 genetically unsolved. According to Nassisi et al., there are two basic explanations for this performance’s relative poorness: (1) because the whole gene was not scanned, the second allele may be located in the gene’s promoter, untranslated regions (UTRs), or another deep intronic region. Additionally, the phenotype may be caused by unrecognized copy number variations (CNVs) in exonic or intronic areas [39]. Hence, sequencing of *ABCA4* locus could genetically resolve this case [37]. (2) As there are multiple phenocopies associated with STGD, it may be necessary to examine the exome or genome of the patient to identify the existence of pathogenic mutations in other genes, possibly additional genes not previously linked with IRDs [37].

The *ABCA4* locus provides insight into a multitude of founder alleles in a particular geographical region. What makes the *ABCA4* locus particularly intriguing is; (1) almost every population has its intrinsic mutations, (2) the wide variance in pathogenic mutations seen among populations of distinct racial and ethnic origins [11]. The most prevalent disease-causing mutation in the *ABCA4* gene, namely c.5882G>A; p.(Gly1961Glu), exhibits a likely origin in Eastern Africa, with frequencies ranging from approximately 8% to 10% in populations hailing from Somalia, Kenya, and Ethiopia. [11, 40]. The dispersal of c.5882G>A; p.(Gly1961Glu) worldwide can be attributed to the migratory movement of populations across different regions. Yet, the allele frequency has declined substantially with evolution, reaching about 0.4% in European nations, implying that the variation is causative across almost all populations [11].

Herein, we report the first detection of the mutation c.5882G>A p.(Gly1961Glu) in the Lebanese population. The presence of the Gly1961Glu allele (homozygous or compound heterozygous) is associated with markedly different phenotypes [11]. In this study, Gly1961Glu was detected in about 30% of our group (2 patients out of 7) in both homozygosity and compound heterozygosity states. Mutational analysis revealed p.Gly1961Glu in a homozygous state in the index of family 31 diagnosed with RCD, while it was in compound heterozygous state along with c.4383G>A; p.(Trp1461Ter) in the patient of family 56. Initially, it was thought that Gly1961Glu was not likely to be pathogenic, mainly when found in the homozygous state [41]. A homozygous c.5882G>A; p.(Gly1961Glu) mutation was described in an asymptomatic 25 years old Somali male with normal vision assessment [40, 41]. However, this was justified by a study in which patients homozygous for c.5882G>A; p.(Gly1961Glu) were reported to have later disease onset (>25 years old) than would be seen in STGD in typical cases [40]. Hence, it is likely to find an individual with normal vision assessment at 25 but may develop symptoms in later life stages [40]. A prior investigation found an association between the c.5882G>A; p.(Gly1961Glu) mutation, with the development of bull’s eye maculopathy and early disruption of central photoreceptors, supporting the notion that this mutant allele has a disease-causing effect [42]. The missense mutation allele c.5882G>A; p.(Gly1961Glu) has been observed to be linked with retinal impairment that is localized to the macula, without being widespread [42]. This glycine-to-glutamine substitution mutation in exon 42 was envisioned to be outside the functional domains of *ABCA4* [43]. c.5882G>A; p.(Gly1961Glu) is expected to affect protein function by reducing ATP binding and ATPase activity, as shown by indirect functional testing [44, 45]. Generally, c.5882G>A; p.(Gly1961Glu) was reported to be cause milder phenotypes. However, it may associate with phenotypes of varying severity [42]. Its actual clinical manifestation may rely on the severity of the other paired mutant allele, as revealed by previous genotype-phenotype investigations [42, 46]. Hence, the type of the combined *ABCA4* mutant alleles in compound heterozygosity determines the phenotype severity, including the age of onset and functional effects [42, 47]. Interestingly, another modifier genetic and external environmental factors that are still unknown [42, 43]. Our index F31:I31 demonstrated a RCD disease pattern, similar to what Burke et al. reported in a Somali patient with a homozygous p.Gly1961Glu mutation, showing the diversity of phenotypes caused by *ABCA4* mutations [40]. Interestingly, Burke et al. have examined 12 individuals with homozygous p.Gly1961Glu and found that all of them have *ABCA4* retinopathies, with severe phenotypes consistent with the existence of additional (modifier) *ABCA4* mutations [40].

In family 56, the mutation M8: c.4383G>A; p.(Trp1461Ter) found with p.(Gly1961Glu) was confirmed through co-segregation analyses: the index presented it in a compound heterozygous state, while the unaffected parents, F56:M56 and F56:F56, were heterozygous carriers of M5 and M8, respectively. This finding provides confirmation of the autosomal recessive inheritance pattern observed in F56:I56. This mutation was found before along with another allele in a patient diagnosed with CRD [34].

M3: c.574G>A; p.(Ala192Thr) and M4: c.1927G>A; p.(Val643Met) detected by NGS in patient F9:I9.1 of family 9 as compound heterozygous were further confirmed by Sanger sequencing. This patient was diagnosed with CRD. Co-segregation analysis was established where M3 and M4 were affirmed by Sanger sequencing in the mother and the father, respectively, validating the autosomal recessive inheritance fashion of CRD in F9:I9.1. Consistent with our results, M3: c.574G>A causing alanine to threonine p.(Ala192Thr) substitution in *ABCA4* was reported previously in a subject with CRD [27]. Moreover, Webster et al. have also found this mutation in an individual clinically diagnosed with STGD [48]. Both CRD and STGD have clinical features in common, such as age of onset, macula involvement, and a progressive decline in visual acuity while retaining relatively preserved night vision [27]. However, unlike STGD, which is restricted to the posterior pole, CRD generally affects the whole retina [27]. The observation of such a common mutation between CRD and STGD shows overlapping between these two disorders [27]. STGD patients can develop substantial, widespread pigmentary degenerative impairments that are comparable to those seen in CRD patients [27]. Additionally, the other mutation M4: c.1927G>A; p.(Val643Met) has also been reported previously by Briggs et al. as a heterozygous mutation with another allele and found to cause STGD [30]. Maugeri et al. have suggested a genotype-phenotype correlation model which demonstrated an opposite relation between the *ABCA4* residual activity and the level of severity of the IRD [49]. According to this model, compound heterozygosity for two severe (null) *ABCA4* mutations cause RCD, the most severe phenotype [49]. Whereas, in case of partial retention of *ABCA4* activity, CRD will result due to compound heterozygosity of a severe and moderately severe mutation, while STGD macular degeneration will appear if a severe mutation is inherited along with a mild *ABCA4* mutation [49].

Family 37 presented a consanguinity case with a child diagnosed with CRD. The homozygous nonsense mutation revealed in the index of family 37, M7: c.6088C>T, causing a stop codon at Arg2030, is likely to destabilize the messenger RNA by the nonsense-mediated decay mechanism in case the protein is expressed because the affected arginine residue at position 2030 is situated in the second nucleotide-binding domain of ABCA4 protein [50, 51]. This mutation has been identified before in heterozygous and compound heterozygous states and associated with STGD or CRD [43, 52, 53]. The patient of family 41, F41:I41, whose parents were relatives, was diagnosed with CRD and was shown to harbor the homozygous missense mutation c.970T>C; p.(Cys324Arg). Interestingly, this mutation was only seen once in a compound heterozygous state with c.4316G>T; p.(Gly1439Val) in a Chinese patient with STGD [33]. Similarly, a homozygous missense mutation in exon 22 of *ABCA4*: c.3259G>A; p.(Glu1087Lys) was associated with RCD in the index patient of family 58. This mutation was previously found in the compound heterozygous state associated with STGD and CRD [54, 55].

In conclusion, eight mutations in the *ABCA4* gene were detected in seven Lebanese patients diagnosed with different forms of related retinopathies. One mutation turned out to be novel; [M1]; c.4330G>C; p.(Trp1408Cys). When Combined with the phenotypic data, our findings show the significant allelic heterogeneity of *the ABCA4* gene. The expanded capabilities of genetic screening, assisted by the utilization of high-resolution diagnostic imaging technologies, have enlarged the phenotypic expression spectrum of *ABCA4-associated* diseases. A thorough understanding of the *ABCA4* mutations and its correlations with the phenotype are indispensable to comprehend its association with different forms of IRDs.

## Data Availability

All data produced in the present work are contained in the manuscript

## Acknowledgments

We express our gratitude to the participants and their families for their dedicated time and efforts in participating in this study. Additionally, we extend our appreciation to 3billion® (Seoul, Korea) for generously providing free whole-exome sequencing to the index patients F37:I37 and F56:I56.

## Author Contributions

SES framed the methodology. AA, CH, and SES conducted the formal analysis. LJ, SES, HA, and MI, LJ and SES investigated. AA, CH, and LJ collected resources. MI and LJ wrote the original draft preparation. SES wrote the review and editing. SES administered the project. AA and SES funded the acquisition.

## Confidentiality

The patient and the family IDs are not known to anyone outside the PI: SES.

